# Shingles vaccination reduces risk of Parkinson’s disease

**DOI:** 10.1101/2022.07.18.22277767

**Authors:** Steven Lehrer, Peter H. Rheinstein

**Author notes:** correspondence to Dr. Steven Lehrer, Box 1236 Radiation Oncology, Mount Sinai Medical Center, 1 Gustave L. Levy Place, New York 10029 or. Data sources described in article all publicly available. Dr. Lehrer and Dr. Rheinstein contributed equally to the conception, writing, and data analysis of this study.

## Abstract

**Background:** Shingles vaccination protects against Alzheimer’s disease (AD), which is related to herpes virus infection. In the current analysis we attempted to determine if herpes zoster vaccination might reduce the risk of PD.

**Methods:** Data on PD prevalence by US state is from Mantri et al, Table 1. They identified 27,538,023 Medicare beneficiaries that met inclusion criteria, of whom 392,214 had a PD diagnosis in 2014. Data on Shingles vaccination among adults aged 60 and over, United States, 2018 is from Terlizzi and Black, Figure 4. The NHIS data from 2008 to 2018 were used for this investigation. The NHIS is a household survey of the civilian, non-institutionalized U.S. population that is conducted nationally. It is continually carried out by the National Center for Health Statistics during the entire year (NCHS). Although follow-ups to completed interviews may be made over the phone, interviews are conducted in respondents’ homes.

**Results:** States with the most PD (lowest age adjusted prevalence ranks) had the lowest proportion of adults aged 60 and over who had ever received a shingles vaccine. Increased vaccination proportion led to significantly reduced female PD prevalence. Increased proportion of Medicare-Medicaid dual eligibility and increased health care spending were associated with diminished proportion of adults who had ever received a shingles vaccine.

**Conclusion:** Shingles vaccination is a wise, cost-effective public health measure that may reduce PD and AD prevalence.

---

The most prevalent age-related movement condition, Parkinson’s disease (PD), is characterized by bradykinesia, resting tremor, unbalanced gait, muscular rigidity, postural instability, as well as some nonmotor symptoms like autonomic and cognitive dysfunctions. Lewy bodies and Lewy neurites collect alpha-synuclein in PD. Select peripheral autonomic nervous system neurons and central nervous system neurons are affected. The incidence of PD rises with age, and the lifetime frequency is 1 percent to 5 percent. Multiple twin studies support the idea that environmental factors play a larger role in disease pathogenesis than do genetic factors, even though evidence for a few rare genetic mutations in a small subset of young people with PD cases offer some insights into the pathogenesis [1].

Although the precise cause of PD is uncertain, mounting evidence points to viral infection as a potential factor. For instance, the varicella zoster virus (VZV) may remain dormant in the ganglia and reawaken because of weakened immunity or aging. Herpes zoster (shingles) is a VZV infection that causes a painful skin rash and blisters on the dermatome infected. Herpes zoster may be related to Parkinson’s disease, according to recent research [2, 3].

Herpes zoster vaccination protects against Alzheimer’s disease (AD), which is related to herpes virus infection [4-7]. In the current analysis we attempted to determine if herpes zoster vaccination might reduce the risk of PD.

## Methods

Data on PD prevalence by US state is from Mantri et al, Table 1 [8]. They identified 27,538,023 Medicare beneficiaries that met inclusion criteria, of whom 392,214 had a PD diagnosis in 2014.

Data on Shingles vaccination among adults aged 60 and over, United States, 2018 is from Terlizzi and Black, Figure 4 [9]. The NHIS data from 2008 to 2018 were used for this investigation. The NHIS is a household survey of the civilian, non-institutionalized U.S. population that is conducted nationally. It is continually carried out by the National Center for Health Statistics during the entire year (NCHS). Although follow-ups to completed interviews may be made over the phone, interviews are conducted in respondents’ homes.

Statistical analysis was performed with SPSS v26 (IBM, Armonk NY).

## Results

1) District of Columbia, 2) New York, 3) Illinois, 4) Connecticut and 5) Florida had the lowest age adjusted prevalence ranks and highest age adjusted prevalence of PD.

Figure 1 shows age adjusted PD prevalence ranks, 50 US states and the District of Columbia, versus proportion of adults who had ever received a shingles vaccine. The relationship is significant (p = 0.005, two tailed). States with the most PD (lowest age adjusted prevalence ranks) had the lowest proportion of adults aged 60 and over who had ever received a shingles vaccine.

**Figure 1.**
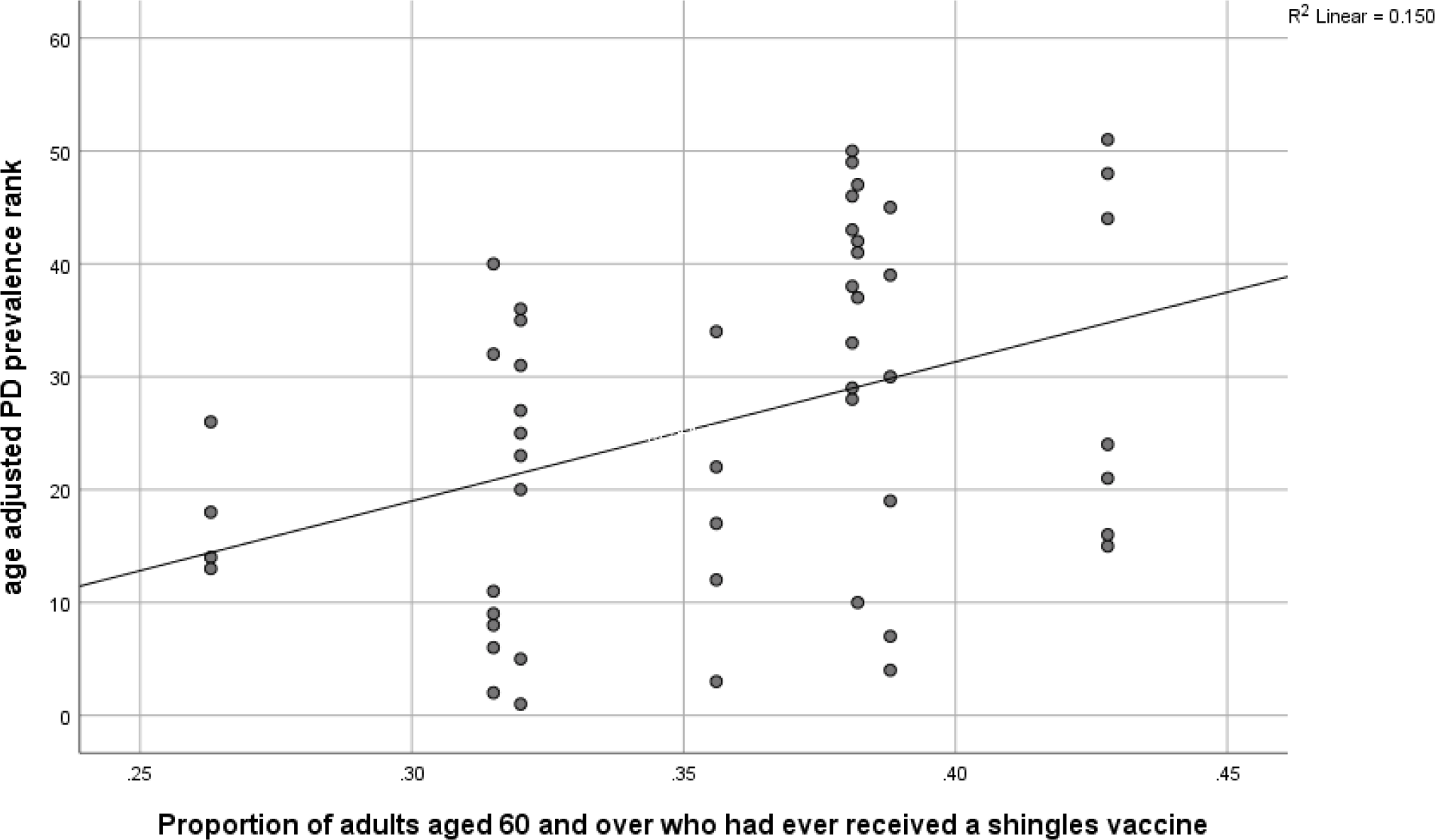
Age adjusted PD prevalence ranks, 50 US states and the District of Columbia, versus proportion of adults who had ever received a shingles vaccine. The relationship is significant (p = 0.005, two tailed). States with the most PD (lowest age adjusted prevalence ranks) had the lowest proportion of adults aged 60 and over who had ever received a shingles vaccine.

Proportion female subjects versus proportion of adults who had ever received a shingles vaccine is in Figure 2. The relationship is significant (p < 0.001, two tailed). Increased vaccination proportion led to significantly reduced female PD prevalence. Men have a higher incidence of PD [10].

**Figure 2.**
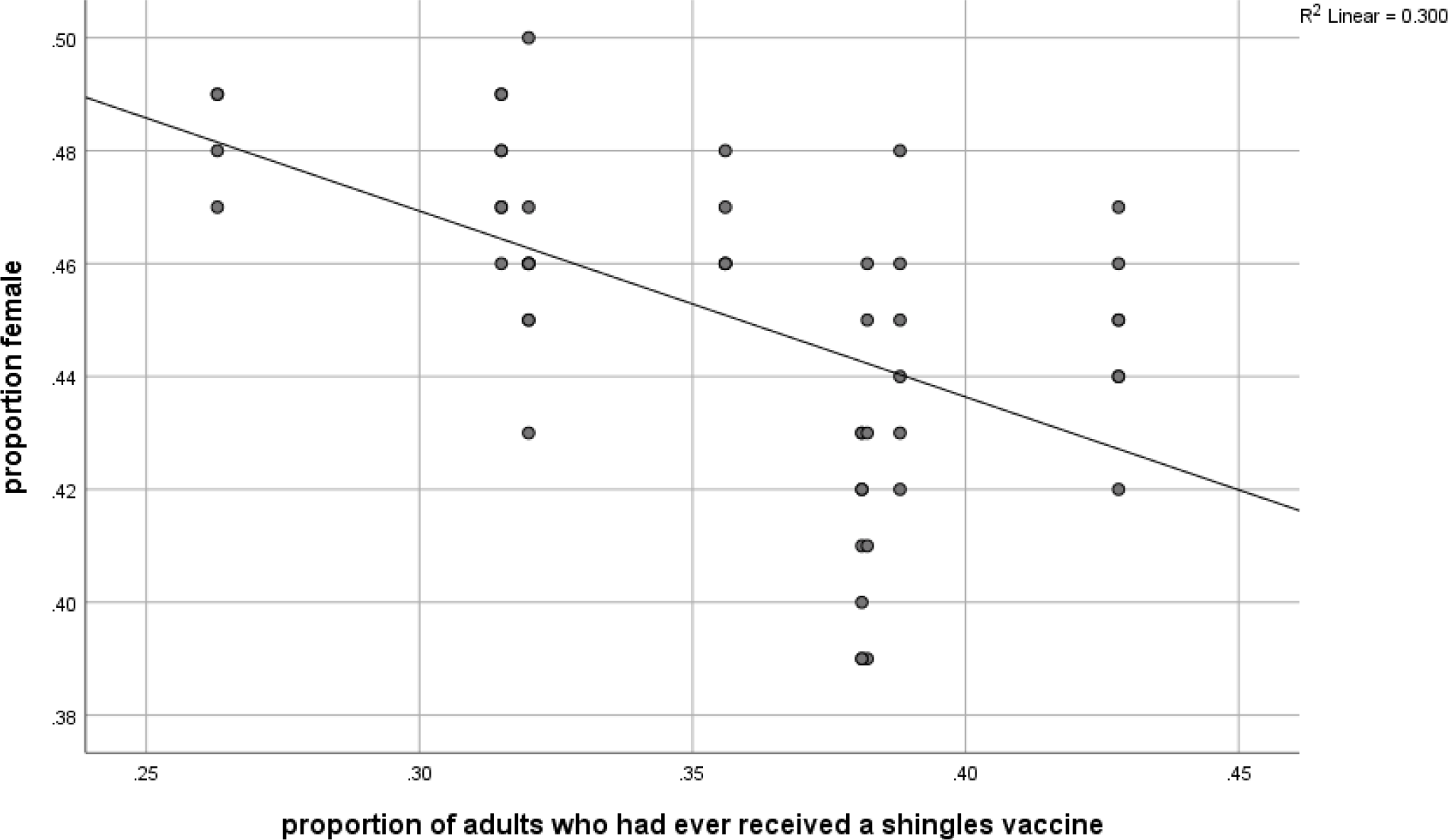
Proportion female subjects versus proportion of adults who had ever received a shingles vaccine. The relationship is significant (p < 0.001, two tailed). Increased vaccination proportion led to significantly reduced female PD prevalence.

Proportion dual eligibility for Medicare and Medicaid versus proportion of adults who had ever received a shingles vaccine is in Figure 3. The relationship is significant (p = 0.003, two tailed). Increased proportion of dual eligibility and increased health care spending were associated with diminished proportion of adults who had ever received a shingles vaccine.

**Figure 3.**
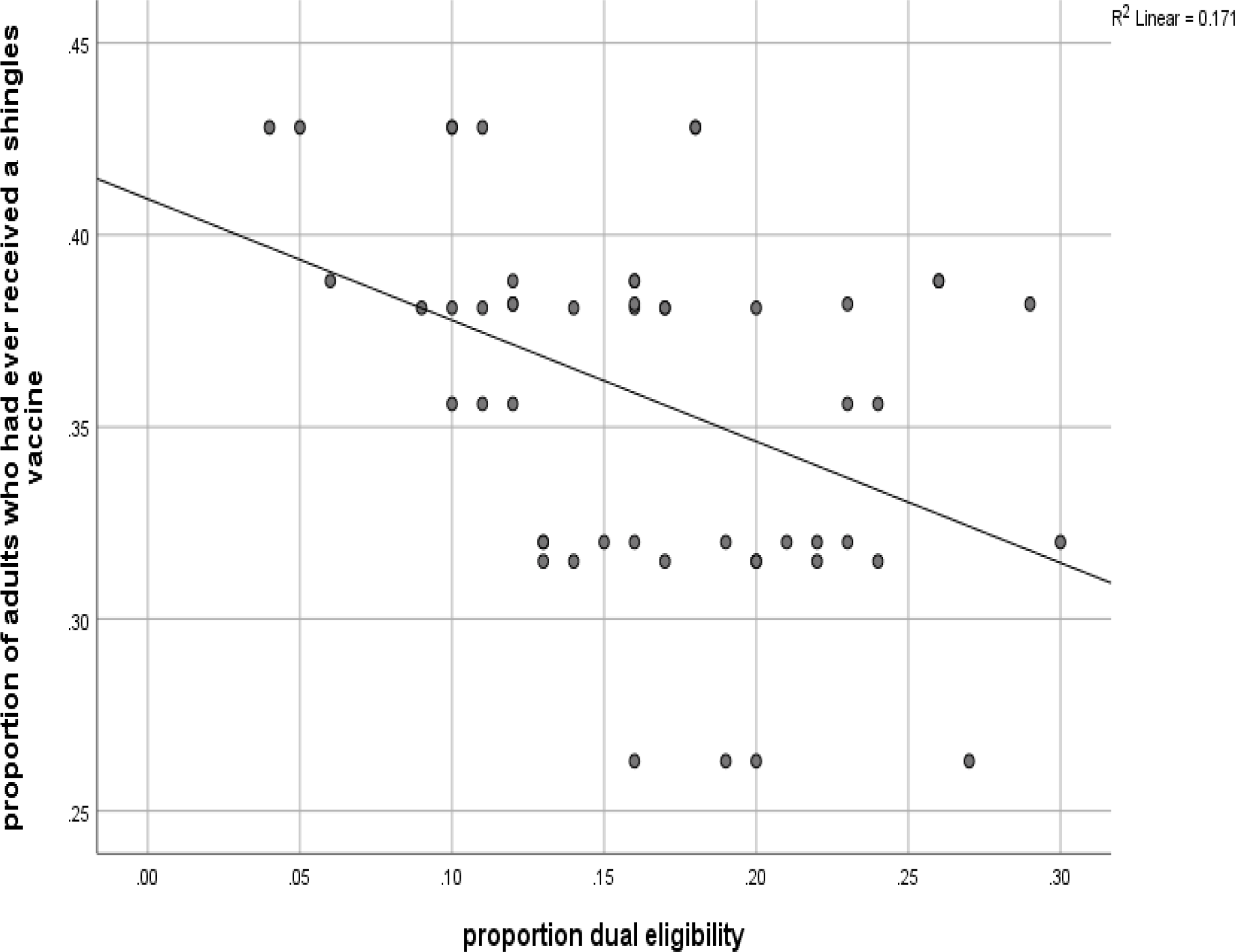
Proportion dual eligibility for Medicare and Medicaid versus proportion of adults who had ever received a shingles vaccine. The relationship is significant (p = 0.003, two tailed). Increased proportion of dual eligibility and increased health care spending were associated with diminished proportion of adults who had ever received a shingles vaccine.

## Discussion

Studies of HZV infection and PD have been unclear as to whether HZV and PD are related. Peripheral T lymphocytes and B lymphocytes may decline over the course of PD. Herpes zoster may have a potential to develop later throughout the normal course of PD because of the reduction in cell-mediated immunity according to a theory put forth by Lai et al [3]

In contrast, Cheng et al report that people with herpes zoster have a higher chance of developing PD and hypothesize that risk may be influenced by the inflammation associated with the herpes zoster infection. To pinpoint the precise pathophysiological relationship between herpes zoster and the risk of PD, more research is necessary [1].

Both AD and PD may be due to late life reactivation of embryologic pathways and processes silenced at birth. Viruses such a HZV may play a part [7].

The ecological fallacy, also known as the ecological inference fallacy, is a logical fallacy in the interpretation of statistical data where inferences about the nature of individuals are derived from inference for the group to which those individuals belong [11]. The fallacy could confound our study’s results. In this instance, rather than from the individuals themselves, assumptions regarding PD in individuals are being made based on the features of the US states in which they reside.

Increased proportion of Medicare Medicaid dual eligibility and increased health care spending associated with diminished proportion of adults who had ever received a shingles vaccine (Figure 3) imply that shingles vaccination is a wise, cost-effective public health measure. Zostavax, the live vaccine, was discontinued in the U.S., November 18, 2020. Shingrix, which is more effective, is given as two intramuscular doses 6 months apart.

Cigarette smoking and coffee drinking reduce risk of PD. Shingles vaccination could be another factor that reduces risk. Further studies are warranted [12].

## Data Availability

All data publicly available.

https://stacks.cdc.gov/view/cdc/90518

